# Structured Expert Judgement Approach of the Health Impact of Various Chemicals and Classes of Chemicals

**DOI:** 10.1101/2024.01.30.24301863

**Authors:** Deniz Marti, David Hanrahan, Ernesto Sanchez-Triana, Mona Wells, Lilian Corra, Howard Hu, Patrick N. Breysse, Amalia Laborde, Jack Caravanos, Roberto Bertollini, Kate Porterfield, Richard Fuller

## Abstract

**Introduction:** Chemical contamination and pollution are an ongoing threat to human health and the environment. The concern over the consequences of chemical exposures at the global level continues to grow. Because resources are constrained, there is a need to prioritize interventions focused on the greatest health impact. Data, especially related to chemical exposures, are rarely available for most substances of concern, and alternate methods to evaluate their impact are needed.

**Structured Expert Judgment (SEJ) Process:** A Structured Expert Judgment^3^ process was performed to provide plausible estimates of health impacts for 16 commonly found pollutants: asbestos, arsenic, benzene, chromium, cadmium, dioxins, fluoride, highly hazardous pesticides (HHPs), lead, mercury, polycyclic-aromatic hydrocarbons (PAHs), polychlorinated biphenyls (PCBs), Per- and Polyfluorinated Substances (PFAs), phthalates, endocrine disrupting chemicals (EDCs), and brominated flame retardants (BRFs). This process, undertaken by sector experts, weighed individual estimations of the probable global health scale health impacts of each pollutant using objective estimates of the expert opinions’ statistical accuracy and informativeness.

**Main Findings:** The foremost substances, in terms of mean projected annual total deaths, were lead, asbestos, arsenic, and HHPs. Lead surpasses the others by a large margin, with an estimated median value of 1.7 million deaths annually. The three other substances averaged between 136,000 and 274,000 deaths per year. Of the 12 other chemicals evaluated, none reached an estimated annual death count exceeding 100,000. These findings underscore the importance of prioritizing available resources on reducing and remediating the impacts of these key pollutants.

**Range of Health Impacts:** Based on the evidence available, experts concluded some of the more notorious chemical pollutants, such as PCBs and dioxin, do not result in high levels of human health impact from a global scale perspective. However, the chemical toxicity of some compounds released in recent decades, such as Endocrine Disrupters and PFAs, cannot be ignored, even if current impacts are limited. Moreover, the impact of some chemicals may be disproportionately large in some geographic areas. Continued research and monitoring are essential; and a preventative approach is needed for chemicals.

**Future Directions:** These results, and potential similar analyses of other chemicals, are provided as inputs to ongoing discussions about priority setting for global chemicals and pollution management. Furthermore, we suggest that this SEJ process be repeated periodically as new information becomes available.

## Introduction

### Chemical Pollution and Health

Chemical contamination and pollution are growing threats to human health and the environment.^1^ In a pivotal acknowledgement, the United Nations Environment Programme (UNEP) recognized chemical pollution as a planetary crisis in 2021, placing it on a par with climate change and biodiversity loss.^2^ This was followed by the decision of the United Nations Environment Assembly in March 2022 that a Science-Policy Panel (SPP) should be established to contribute further to the sound management of chemicals and waste and to prevent pollution.^3^

There are many potentially harmful chemicals already in use, and new chemicals are developed daily.^4^ In 2020, the World Health Organization (WHO) published a list of the ‘Top Ten’ chemicals of public health concern.^5^ A great deal of work has been done to evaluate and mitigate the hazards and risks of individual chemicals by regulatory bodies in multiple jurisdictions.^6,7^ Such research assesses the potential (hazard) and likelihood (risk) that harm might occur as a result of exposure to a chemical. However, the most important factor in terms of threats and setting priority for action is actual impact on the public. Impact describes the harm that is actually occurring, and can be estimated based on a combination of risk and exposure analysis.^8^ While impact is often calculated in discrete circumstances, there is a lack of reliable and comparable information on large scale impacts.

The best available data on the overall health impacts of chemicals comes from the Global Burden of Disease (GBD). This database includes several chemicals as risk factors for disease and their global impacts, based on published health data.^9^ However, coverage of chemical pollutants is limited because the necessary exposure data for many of the toxins are simply unavailable. Only occupational exposures are available for most chemicals, except lead. Therefore, an SEJ process can effectively address this limitation and help overcome the lack of environmental exposure data for chemicals.^10^ SEJ has been successfully applied in a range of relevant fields.^11^

### Information Gaps and Value of SEJ Process

In order to address information gaps and to provide plausible estimates of global scale health impacts for 16 commonly occurring pollutants, the SEJ process was used.

SEJ is a systematic and rigorous method for collecting and combining expert elicitations about the information that is hard or impossible to obtain. It distinguishes from other expert elicitation approaches in the sense that it incorporates a weighing system based on experts’ performances. The aggregation of expert elicitation involves assigning differentiated weights to each expert’s contribution, where these weights are calibrated based on the individual performance of the experts. Such performance is assessed with respect to their statistical accuracy and informativeness. First experts are asked to provide their best estimates for the variables (i.e., calibration variables) within their expertise, for which true values are known. Based on their accuracy in their estimates for those calibration variables, this procedure results in weights for each expert. These performance-based weights that are derived from calibration processes then are used in generating a probability distribution. This final combined elicitation is called decision maker (DM).

SEJ was selected to support the prioritization of chemical pollutants because of the lack of comprehensive and reliable data on exposure and health impacts of many pollutants, despite the widespread use of the chemicals and frequent (more or less reliable) accounts of damage or harm. Much good research and analysis continues on these pollutants but the complexity of the effects and the slow pace of peer-reviewed science means that plausible expert opinion has an important role to play in helping to set priorities for investigation and intervention.

### Methodology and Implementation of SEJ Process

The SEJ process involved nine experts who provided individual estimations of four numerical variables relating to death and disability impacts of each of 16 chemicals. This set of chemicals included metals and classes of chemicals as well as individual compounds. The selection of the chemicals to be assessed was based on a range of sources including WHO’s “Chemicals of Public Health Concerns;”^5^ relevant UN conventions;^12,13,14^ and discussions among the project team. The experts were identified through discussions with recognized and respected researchers in these fields and they were then invited to participate. In the end, the number of nine experts who completed the whole process was close to the design ideal of ten contributors.

In the initial stage, each expert was asked separately to provide answers to a set of 17 calibration questions from the broad field of chemicals, pollution and health. These answers were subsequently used to weigh the study findings towards those respondents who provided the most accurate responses at calibration (details of the calibration and elicitation processes are provided in the Supplementary Materials).

In the following elicitation phase, experts were requested individually to provide their best estimates (at 5%, 50%, and 95% percentiles, see Supplementary Material for more detail) of premature deaths and Disability-Adjusted Life Years (DALYs) Lost (in year 2019) resulting from each of the 16 chemicals. The experts were also asked what percentage occurred in High-Income Countries (HICs) and in Low- and Middle-Income countries (LMICs). They were asked to suggest which countries were the most impacted (both LMICs and HICs) and whether the existing levels of science regarding dose-response relations and exposure levels were adequate.

The individual responses by each of the experts to all of the questions were then analyzed, taking into account the weightings derived from the calibration, and the integrated results presented (further details can be found in the Supplementary Materials).

### Key Findings

The key findings can be summarized in the following charts and tables.

**Fig 1.**
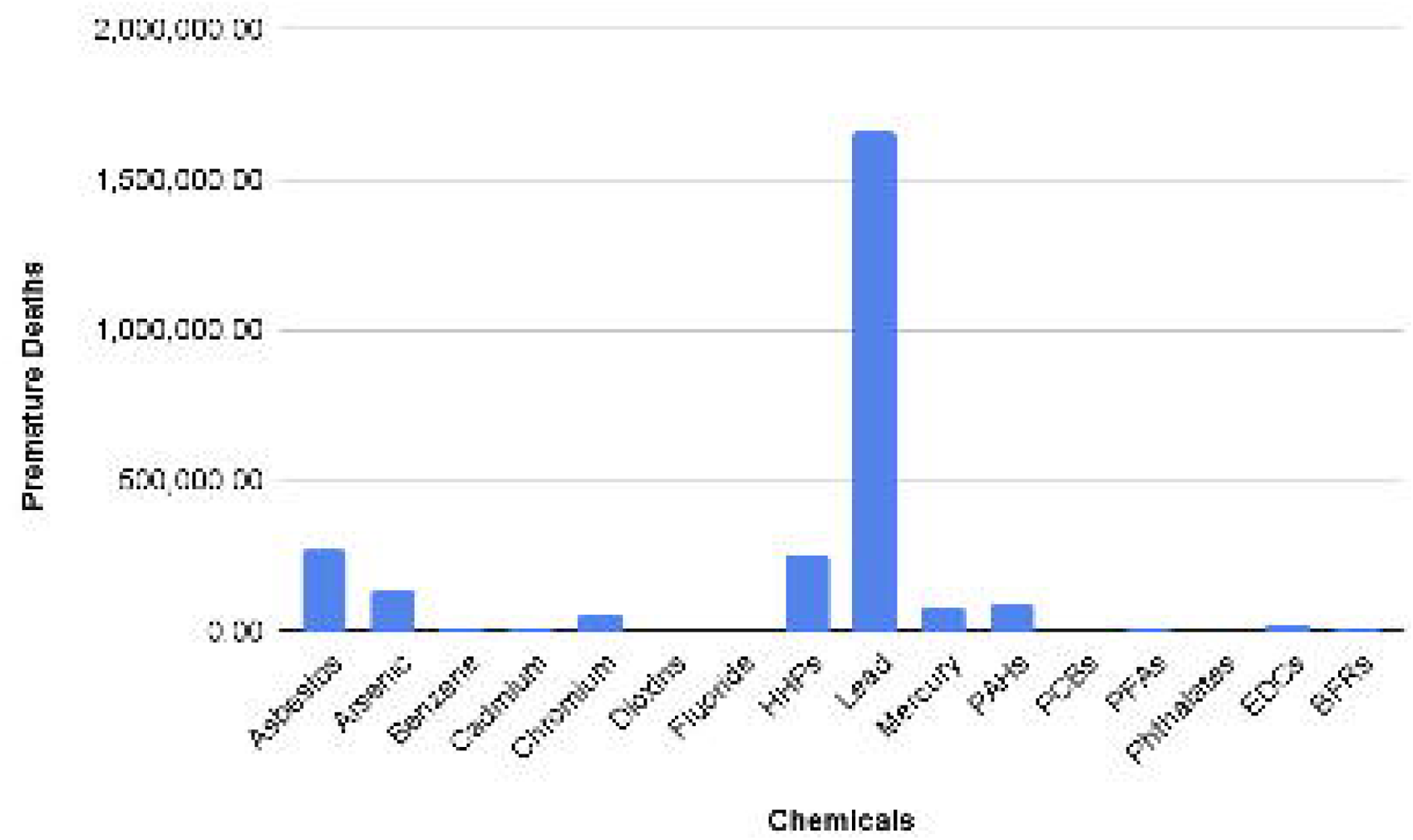
Premature Deaths, Performance Weighted (PW) 50% Decision Marker (DM):

Figure 1 provides a visual representation of the estimated number of premature deaths associated with exposure to various environmental chemicals based on the performance-weighted decision maker’s 50% percentile. The y-axis quantifies the number of premature deaths, while the x-axis lists the chemicals analyzed. As shown in the figure, Lead exposure remarkably accounts for the highest estimated number of premature deaths, overshadowing the impact of the other chemicals significantly. The subsequent levels of premature deaths associated with Asbestos exposure, although substantially lower than Lead. The remaining chemicals contribute to a lesser but non-negligible number of premature deaths.The tabulated form of the results is as follows:

**Table 1.** Premature Deaths (PW) by Chemical.

| Chemical | PW5% | PW50% | PW95% |
| --- | --- | --- | --- |
| Asbestos | 48,200 | 274,000 | 346,000 |
| Arsenic | 22 | 136,000 | 482,000 |
| Benzene | 2 | 3,472 | 6,998 |
| Cadmium | 3 | 8,327 | 48,900 |
| Chromium | 4 | 59,000 | 146,000 |
| Dioxins | 0 | 48 | 326 |
| Fluoride | 0 | 48 | 138 |
| HHPs | 142 | 252,000 | 484,000 |
| Lead | 945 | 1,660,000 | 3,230,000 |
| Mercury | 1 | 79,800 | 150,000 |
| PAHs | 9 | 89,900 | 148,000 |
| PCBs | 0 | 25 | 1,989 |
| PFAs | 0 | 9,460 | 50,000 |
| Phthalates | 0 | 10 | 14,700 |
| EDCs | 0 | 23,900 | 452,000 |
| BFRs | 0 | 8,591 | 48,600 |

*Note: Figures have been rounded to the nearest whole number*

### Findings by Chemical

All estimates are performance-weighted estimates.

**Fig 2.**
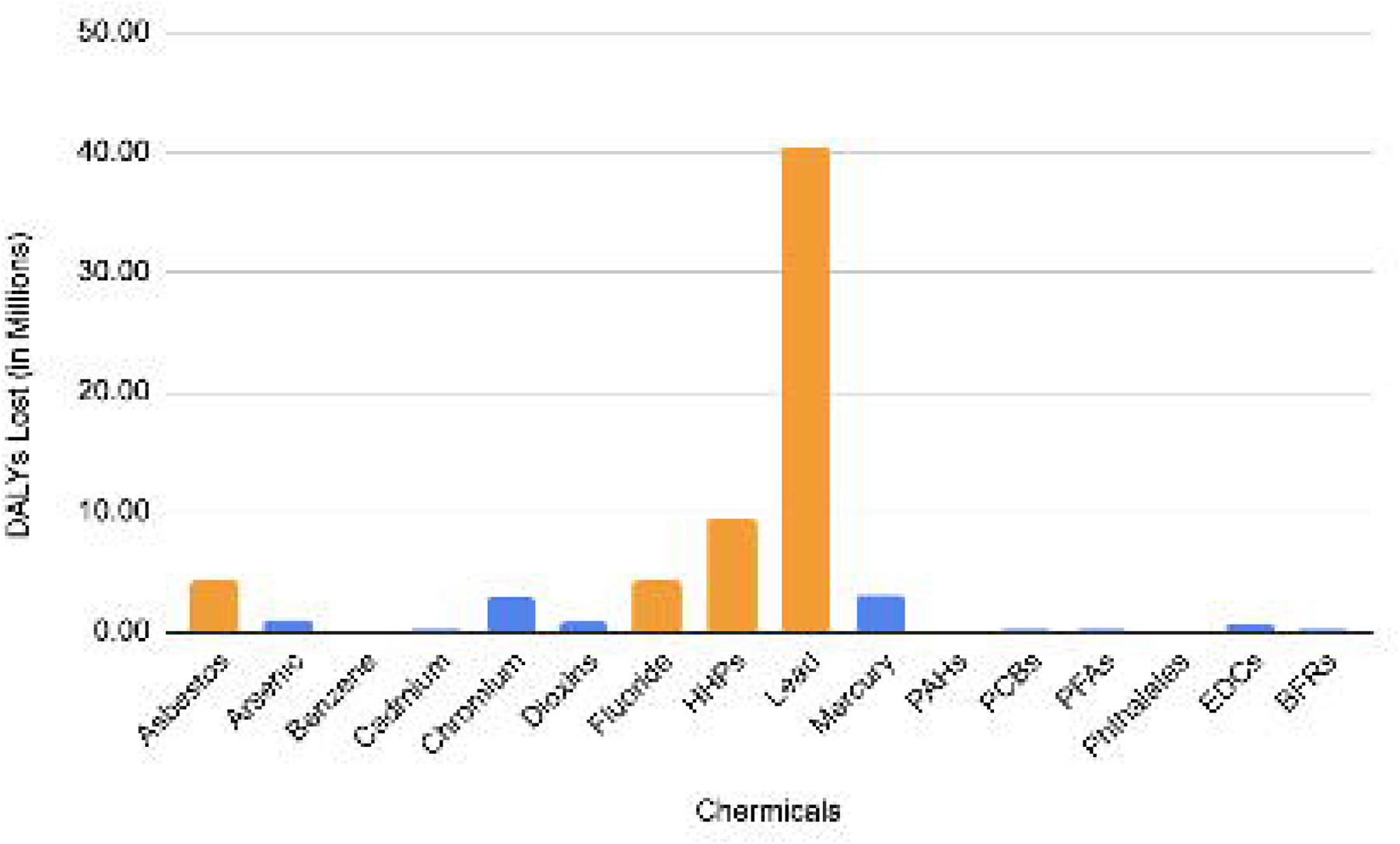
The Impact of Different Chemical Exposures on Disability-Adjusted Life Years (50% Years) (DALYs)

Figure 2 illustrates the health impact of various chemicals in terms of median DALYs. Lead shows the highest impact, with a total of 40,500,000 DALYs. This is followed by HHPs, contributing to 9,460,000 DALYs. Additionally, the graph includes the DALYs associated with Fluoride and Asbestos, which are 4,320,000 and 4,230,000 respectively. These data provide a comparative overview of the health burdens posed by each chemical, highlighting the significant disparities in their impact.

#### Asbestos (ASB)

While banned in most HIC, asbestos is still mined and commonly used in many LICs as a building material and used with little personal protection.^16^ An estimate of 274,000 premature deaths, and 4,230,000 lost DALYs, are attributed to asbestos annually. Of these deaths, experts estimate 57% occur in HICs, and 43% in LMICs. 43% of experts agree that the current level of science regarding asbestos dose-response analysis is adequate; however, only 50% agree that the current level of science regarding asbestos exposure levels is adequate. Experts noted that there is inadequate data on exposures in LMICs, making the burden of disease potentially greater than current statistics indicate.

#### Arsenic (AS)

Arsenic is naturally present at high levels in the groundwater of several countries, and is used industrially as an alloying agent.^17^ An estimate of 136,000 premature deaths, and 987,000 lost DALYs, are attributed to arsenic poisoning annually. Of these deaths, experts estimate 12% occur in HICs, and 88% in LMICs. 66% of experts agree that the current level of science regarding arsenic dose-response analysis is adequate; however, 66% agree that the current level of science regarding arsenic exposure levels is inadequate, especially for LMICs. Additionally, experts stated a need for further research on arsenic’s impact on cardiovascular disease.

#### Benzene (BZ)

Benzene is produced through natural and human processes, and is used to produce lubricants, rubbers, dyes, and pesticides.^18^ An estimate of 3,472 premature deaths, and 98,800 lost DALYs, are attributed to benzene annually. Of these deaths, experts estimate 31% occur in HICs, and 69% in LMICs. 100% of experts agree that the current level of science regarding benzene dose-response analysis is adequate; however, 80% agree that the current level of science regarding benzene exposure levels is inadequate.

#### Cadmium (CD)

Cadmium’s primary use is in the production of batteries, alloys, coatings, and solar panels.^19^ An estimate of 8,372 premature deaths, and 194,000 lost DALYs, are attributed to cadmium poisoning annually. Of these deaths, experts estimate 2% occur in HICs, and 98% in LMICs. 60% of experts agree that the current level of science regarding cadmium dose-response analysis is inadequate; and 80% agree that the current level of science regarding cadmium exposure levels is inadequate. Experts noted that there are inadequate data on cadmium exposures in LMICs, making the burden of disease potentially greater than current statistics indicate. Experts additionally stated that while the global production of cadmium at the present is relatively flat, future battery use may change this, and subsequent exposure rates.

#### Chromium Hex (CR)

Chromium is largely used in the production of stainless steel and several alloys.^20^ An estimate of 59,000 premature deaths, and 2,930,000 lost DALYs, are attributed to hexavalent chromium exposure annually. Of these deaths, experts estimate 8% occur in HICs, and 92% in LMICs. 58% of experts agree that the current level of science regarding chromium dose-response analysis is adequate; and 70% agree that the current level of science regarding chromium exposure levels is inadequate. Experts noted that data about chromium’s potential pathways are lacking, especially for LMICs, making the burden of disease potentially greater than current statistics indicate.

#### Dioxins (DF)

Dioxins are largely by-products of industrial practices, including smelting, chlorine bleaching, and the manufacturing of pesticides.^21^ An estimate of less than 100 premature deaths, and 901,000 lost DALYs, are attributed to dioxins annually. Of these deaths, experts estimate 10% occur in HICs, and 90% in LMICs. 58% of experts agree that the current level of science regarding dioxins dose-response analysis is inadequate; and 71% agree that the current level of science regarding dioxins exposure levels is inadequate.

#### Fluoride (FF)

Fluoride’s primary use is in water fluoridation to promote oral health and prevent tooth decay.^22^ An estimate of less than 100 premature deaths, and 4,320,000 lost DALYs, are attributed to fluoride annually. Of these deaths, experts estimate 15% occur in HICs, and 85% in LMICs. 71% of experts agree that the current level of science regarding fluoride dose-response analysis is inadequate; and, 71% agree that the current level of science regarding fluoride exposure levels is inadequate.

#### Highly Hazardous Pesticides (HHPs)

HHPs are pesticides that cause disproportionate harm to the environment and human health.^23^ An estimate of 252,000 premature deaths, and 9,460,000 lost DALYs, are attributed to HHPs. Of these deaths, experts estimate 15% occur in HICs, and 85% in LMICs. 57% of experts agree that the current level of science regarding HHPs dose-response analysis is adequate; however, 88% agree that the current level of science regarding HHPs exposure levels is inadequate.

#### Lead (PB)

Lead is used in a wide array of products, including paint, ceramics, pipes, cosmetics, spices, and batteries.^24^ An estimate of 1,660,000 premature deaths, and 40,500,000 lost DALYs, are attributed to lead annually. Of these deaths, experts estimate 5% occur in HICs, and 86% in LMICs. 86% of experts agree that the current level of science regarding lead dose-response analysis is adequate; and 71% agree that the current level of science regarding lead exposure levels is adequate.

#### Mercury (HG)

Mercury exists in many forms, and is a significant by-product of small-scale gold mining and other industrial processes.^25^ An estimate of 79,800 premature deaths, and 2,950,000 lost DALYs, are attributed to mercury exposure annually. Of these deaths, experts estimate 11% occur in HICs, and 88% in LMICs. 57% of experts agree that the current level of science regarding mercury dose-response analysis is adequate; however, 71% agree that the current level of science regarding mercury exposure levels is inadequate.

#### Polycyclic Aromatic Hydrocarbons (PAHs)

PAHs are formed by incomplete combustion of coal, oil, and gasoline, and are used in the production of dyes, plastics, and pesticides.^26^ An estimate of 89,900 premature deaths, and 1,848,000 lost DALYs, are attributed to PAHs annually. Of these deaths, experts estimate 11% occur in HICs, and 85% in LMICs. 57% of experts agree that the current level of science regarding PAH dose-response analysis is adequate; however, 57% agree that the current level of science regarding PAH exposure levels is inadequate.

#### Polychlorinated Biphenyls (PCBs)

While the production of PCBs are banned in some countries, including the United States, their longevity allows PCBs to still be used in an array of industrial and commercial applications, including electrical equipment, plasticizers,and pigments.^27^ An estimate of less than 100 premature deaths, and 288,000 lost DALYs, are attributed to PCBs annually. Of these deaths, experts estimate 39% occur in HICs, and 56% in LMICs. 71% of experts agree that the current level of science regarding PCB dose-response analysis is inadequate; and 71% agree that the current level of science regarding PCB exposure levels is inadequate.

#### Polyfluorinated Substances (PFAs)

PFAs are widely used in consumer products including nonstick cookware, water-repellent clothing, and stain-resistant consumer goods.^28^ An estimate of 50,000 premature deaths, and 183,000 lost DALYs, are attributed to PFAs annually. Of these deaths, experts estimate less than one percent occur in HICs, and less than one percent in LMICs. 88% of experts agree that the current level of science regarding PFA dose-response analysis is inadequate; and 88% agree that the current level of science regarding PFA exposure levels is inadequate.

#### Phthalates (FH)

Phthalates are mainly used as plasticizers, added to polyvinyl chloride plastics as a softener.^29^ An estimate of less than 100 premature deaths, and 3,988 lost DALYs, are attributed to phthalates annually. Of these deaths, experts estimate 53% occur in HICs, and 43% in LMICs. 57% of experts agree that the current level of science regarding phthalates dose-response analysis is inadequate; and 100% agree that the current level of science regarding phthalates exposure levels is inadequate.

#### Endocrine Disrupting Chemicals (EDCs)

ECDs are substances found in food, consumer products, as well as the environment, that disrupt hormonal systems.^30^ An estimate of 23,900 premature deaths, and 647,000 lost DALYs, are attributed to EDCs annually. Of these deaths, experts estimate 50% occur in HICs, and 50% in LMICs. 86% of experts agree that the current level of science regarding EDC dose-response analysis is inadequate; and 100% agree that the current level of science regarding EDC exposure levels is inadequate. Experts noted that there is inadequate data on exposures of EDCs, with their ultimate health impacts unclear.

#### Brominated Flame Retardants (BFRs)

BFRs are used as additives to consumer products to reduce fire-related injury and damage.^31^ An estimate of 8,591 premature deaths, and 199,000 lost DALYs, are attributed to BFRs annually. Of these deaths, experts estimate 50% occur in HICs, and 50% in LMICs. 100% of experts agree that the current level of science regarding BFR dose-response analysis is inadequate; and 100% agree that the current level of science regarding BFR exposure levels is inadequate. Experts noted that there is inadequate data on exposures globally, making the burden of disease potentially greater than current statistics indicate.

A summary of the HICs and LMICs considered to be most impacted by each chemical, as well as their most severe pathways of exposure, can be found in the Supplementary Material.

### Main findings

The top four substances identified, in terms of mean estimated annual total deaths, were lead, asbestos, arsenic, and HHPs. Of these, the deaths attributed to lead were the highest by a large margin, at an estimated mean value of 1.7 million annually.

## Discussion

This analysis estimates the impact of these chemicals on a global scale. It is critical to note, however, that these exposures are not equally distributed. As a result, certain communities may experience high exposures to some of the toxicants examined, even though the global estimates of impact are modest or low (e.g.., asbestos in those countries still mining it or using it).^32^ This study should be considered a tool for the prioritization of chemical pollutants only on a macro level, as the findings may not align with the concerns of specific towns and municipalities. Impact at local sites where toxins might cause substantial damage ought to be considered on their local evidence. Further information on countries impacted most by each chemical exposure can be found in the Supplementary Material.

In the absence of extensive empirical data on the impacts of the various substances, the judgements of the expert panel provide a reasonable basis for setting priorities for interventions to tackle the most hazardous chemicals from a global perspective.

A basic finding of the panel was that the most hazardous substances are traditional ones: lead, asbestos and arsenic, along with HHPs. This is clearly due in part to the very widespread use of these substances and their persistence in the environment. The prominence of these substances may reflect familiarity with their hazards, but considerable data exist on the scale of their impact.

Multilateral Environmental Agreements (MEAs) exist for only a few of these chemicals, notably mercury (Minamata Convention)^13^ and HHPs (Stockholm Convention).^14^ Others are the concern of the Global Chemicals Framework, or the Strategic Approach to International Chemicals Management (SAICM).^33^

Some chemicals considered to have high political priority, such as PFAS and Phthalates, were not estimated to be among those chemicals with the highest impacts. Additionally, chemicals that have had considerable past attention (dioxins, PCBs) were similarly estimated as having low impacts. In this context, history has shown that chemicals for which risk is newly established gather a great deal of regulatory attention, and enforcement of these regulations can substantially mitigate exposure and impact. It is also worth noting that while regulatory action is more common in high-income countries, and chemical regulatory enforcement is typically lax in low and middle-income countries,^34^ expert assessments were taken at a global level. Nevertheless, one would expect that strong attention to one particular class of chemicals would flow over into work at a global level.

The impact for lead is much higher than previous estimates from IHME but is considerably lower than a recent Lancet paper, which estimates 5.5 million deaths for 2019,^35^ a datum point that was not provided to the experts as publication post-dated the elicitation process. It is worth noting that lead has only recently resurged as an issue of note, as international response to lead has been minimal since the phase out of lead in gasoline. The burden of disease estimates for lead indicates it to be larger than that of HIV/AIDS, malaria, etc.

The moral here is that old but well-known risks are still the largest we face. They have partly been forgotten as the world has turned its attention to plastics and PFAs. Prioritization of pollution control efforts within the UN system and other international aid efforts ought to be focused on those chemicals with the largest impact, and not only those chemicals that garner the most attention. The significant amount of action by industry and NGO groups on favorite chemicals issues will make this very difficult to put into practice. In view of the current knowledge gaps, it is important not to lose sight of the uncertainties expressed in Table 1. For example EDC’s median value for premature deaths (23,900) does place it among the top risks but its 95% value (432,000) reminds us to keep an eye on it, as its risk level could change as more knowledge is gained.

Despite efforts such as this panel to quantify impacts from hazardous chemicals, many uncertainties persist, which should guide future research requirements. The logic of precautionary approaches to emerging chemicals concerns, and ought to be sound, followed regardless of expert opinion on impact.

## Supporting information

Supplemental Files Zip

## Data Availability

All relevant data are within the manuscript and its Supporting Information files.

## Acknowledgements

Special thanks to Roger Cooke at Resources for the Future and Tina Nane at T.U. Delft for their contributions to this manuscript.

