## Supplemental Files Zip for "Structured Expert Judgement Approach of the Health Impact of Various Chemicals and Classes of Chemicals": SEJ Supplementary Material (1).docx

**SI Structured Expert Judgment for Global Burden of Exposure to Metals:**

**Details on Experts and Scoring**

**Deniz Marti et al**

1. **Classical Model Performance Measures and Combination**
   a. Statistical accuracy
   b. Information
   c. Combination: Decision Maker
2. **Elicitation Results**
   a. Expert scoring
   b. Range graphs
   c. Robustness on items

1. **Classical Model Performance Measures and Combination**

The classical model for Structured expert judgment was introduced in (Cooke, 1991). Out−of −sample validation was studied in Eggstaff et al, 2014^36^ and Colson and Cooke, 2017.^37^ Forecast accuracy and persistence of performance measures were studied in (Cooke et al 2020). This reference also links to the current SEJ database. Comparison with the popular alternative combination of averaging quantiles was the subject of Cooke, 2022.^38^

There are two generic, quantitative measures of performance, *statistical accuracy* (aka *calibration)* and *information*. Loosely, statistical accuracy denotes the statistical likelihood that a set of experimental results correspond, in a statistical sense, with an expert’s assessments. Information measures the degree to which a distribution is concentrated. To simplify the exposition, we assume that the 5%, 50% and 95% values were elicited.

***Statistical accuracy***

Assume that each expert assesses 5, 50 and 95 percentiles for each quantity. Each expert divides the range of possible values into 4 inter-quantile intervals for which his/her probabilities are known, namely *p_1_* = 0.05: less than or equal to the 5% value, = 0.45: greater than the 5% value and less than or equal to the 50% value, etc.

If N quantities are assessed, each expert may be regarded as a statistical hypothesis, namely that each realization falls in one of the four inter-quantile intervals with probability vector.

*p= (0.05, 0.45, 0.45, 0.05).*

Suppose we have realizations *x_1_,…x_N_* of these quantities. We may then form the sample distribution of the expert's inter quantile intervals as:

*s_1_(e) = #{ i | x_i_ ≤ 5% quantile}/N*

*s_2_(e) = #{ i | 5% quantile < x_i_ ≤ 50% quantile}/N*

*s_3_(e) = #{ i | 50% quantile < x_i_ ≤ 95% quantile}/N*

*s_4_(e) = #{ i | 95% quantile < x_i_ }/N*

*s(e) = (s_1_(e),…s_4_(e))*

Note that the sample distribution depends on the expert *e*. If the realizations are indeed drawn independently from a distribution with quantiles as stated by the expert then the quantity

*2NI(s(e)| p) = 2N ∑_i=1..4_ s_i_ ln(s_i_ / p_i_)*	(A1.1)

is asymptotically distributed as a chi-square variable with 3 degrees of freedom. This is the so-called likelihood ratio statistic, and *I(s | p)* is the relative information of distributions with respect to *p*. If we extract the leading term of the logarithm we obtain the familiar chi-square test statistic for goodness of fit. There are advantages in using the form in (A1*.*1).^10^

If after a few realizations the expert were to see that all realization fell outside his 90% central confidence intervals, he might conclude that these intervals were too narrow and might broaden them on subsequent assessments. This means that for this expert the uncertainty distributions are *not* independent, and he learns from the realizations. Expert learning is not a goal of an expert judgment study, and his joint distribution is not elicited. Rather, the decision maker wants experts who do not need to learn from the elicitation. Hence the decision maker scores expert *e* as the statistical likelihood of the hypothesis

*H_e_: "the inter-quantile interval containing the true value for each variable is drawn independently from probability vector p."*

A simple test for this hypothesis uses the test statistic *(A1.1*), and the likelihood, or p-value, or **statistical accuracy score** of this hypothesis, is:

*Sa(e)* = *p-value = Prob{ 2NI(s(e)| p)≥ r | H_e_}*

where *r* is the value of *(A1.1)* based on the observed values *x_1_,…x_N_*. It is the probability under hypothesis H_e_ that a deviation at least as great as *r* should be observed on *N* realizations if *H_e_* were true. Statistical accuracy scores are absolute and can be compared across studies. However, before doing so, it is appropriate to equalize the power of the different hypothesis tests by equalizing the effective number of realizations. To compare scores on two data sets with *N* and *N’ realizations*, we simply use the minimum of *N* and *N'* in *(A1.1*), without changing the sample distribution *s*. In some cases involving multiple realizations of one and the same assessment, the effective number of seed variables is based on the number of assessments and not the number of realizations.

Although the statistical accuracy score uses the language of simple hypothesis testing, it must be emphasized that we are not rejecting expert-hypotheses; rather we are using this language to measure the degree to which the data supports the hypothesis that the expert's probabilities are accurate. Low scores, near zero, mean that it is unlikely that the expert’s probabilities are correct.

***Information***

The second scoring variable is information. Loosely, the information in a distribution is the degree to which the distribution is concentrated. Information cannot be measured absolutely, but only with respect to a background measure. Being concentrated or "spread out" is measured relative to some other distribution.

Measuring information requires associating a density to each variable based on each expert’s quantile assessments. To do this, we use the unique density that complies with the experts' quantiles and is minimally informative with respect to a background measure. This density can easily be found with the method of Lagrange multipliers. For a uniform background measure, the density is constant between the assessed quantiles, and is such that the total mass between the quantiles agrees with *p*. The background measure is not elicited from experts as indeed it must be the same for all experts; instead it is chosen by the analyst.

The uniform and log-uniform background measures require an *intrinsic range* on which these measures are concentrated. The classical model implements the so-called *k*% overshoot rule: for each item we consider the smallest interval *I = [L, U]* containing all the assessed quantiles of all experts and the realization, if known. This interval is extended to

*I^*^ = [L^*^, U^*^]; L^*^ = L – k(U-L)/100; U^*^ = U + k(U-L)/100.*

The value of k is chosen by the analyst. A large value of *k* tends to make all experts look quite informative and tends to suppress the relative differences in information scores. The **information score** of expert *e* on assessments for uncertain quantities 1…N is

*Inf (e) =Average Relative information wrt Background = (1/N) ∑_i = 1..N_ I(f_e,i_ | g_i_)*

where *g_i_* is the background density for variable *i* and *f_e,i_* is expert *e*'s density for item i. This is proportional to the relative information of the expert's joint distribution given the background, under the assumption that the variables are independent. As with statistical accuracy, the assumption of independence here reflects a desideratum of the decision maker and not an elicited feature of the expert's joint distribution. The information score does not depend on the realizations. An expert can give himself a high information score by choosing his quantiles very close together.

Evidently, the information score of *e* depends on the intrinsic range and on the assessments of the other experts. Hence, information scores cannot be compared across studies.

Of course, other measures of concentrated-ness could be contemplated. The above information score is chosen because it is:

- Familiar
- Tail insensitive
- Scale invariant
- Slow

The latter property means that relative information is a slow function; large changes in the expert assessments produce only modest changes in the information score. This contrasts with the likelihood function in the statistical accuracy score, which is a very fast function. This causes the product of statistical accuracy and information to be driven by the statistical accuracy score. It also means that modest changes in informativeness correspond to sizable changes in the distributions. Increasing informativeness by a factor *2* roughly corresponds to halving the distance between the 95 and 5 percentiles.

***Combination: Decision Maker***

The **combined score** of expert *e* will serve as an (unnormalized) weight for *e*:

*w_α_(e) = Sa (e) × Inf (e) × 1_α_(Sa(e) ≥ α),	(A1.2)*

where *1_α_(Sa (e)) = 1* if Sa*(e) ≥ α,* and is zero otherwise. The combined score thus depends on *α*. If Sa*(e)* falls below cut-off level *α* expert *e* is unweighted. The presence of a cut-off level is imposed by the requirement that the combined score be an asymptotically strictly proper scoring rule. That is, an expert maximizes his/her long run expected score by and only by ensuring that his probabilities *p= (0.05, 0.45, 0.45, 0.05)* correspond to his/her true beliefs. *α* is similar to a significance level in simple hypothesis testing, but its origin is indeed different. The goal of scoring is not to “reject” hypotheses, but to measure “goodness” with a strictly proper scoring rule.

A combination of expert assessments is called a "decision maker" (DM). All decision makers discussed here are examples of linear pooling. The classical model is essentially a method for deriving weights in a linear pool. "Good expertise" corresponds to good statistical accuracy (high statistical likelihood, high p-value) and high information. We want weights which reward good expertise and which pass these virtues on to the decision maker.

The reward aspect of weights is very important. We could simply solve the following optimization problem: find a set of weights such that the linear pool under these weights maximizes the product of statistical accuracy and information. Solving this problem on real data, one finds that the weights do not generally reflect the performance of the individual experts. As we do not want an expert's influence on the decision maker to appear haphazard, and we do not want to encourage experts to game the system by tilting their assessments to achieve a desired outcome, we must impose a strictly scoring rule constraint on the weighing scheme.

The scoring rule constraint requires the term *1_α_(statistical accuracy score),* but does not say what value of *α* we should choose. Therefore, we choose *α* so as to maximize the combined score of the resulting decision maker. Let *DM_α_(i)* be the result of linear pooling for item *i* with weights proportional to (*A1.*2):

*DM_α_(i) = ∑_e=1,..E_ w_α_(e) f_e,i_ / ∑_e=1,..E_ w_α_(e)*	(*A1.*3)

The *optimized global weight DM* is *DM_α*_* where *α** maximizes

*statistical accuracy score(DM_a_) × information score(DM_α_).*	(*A1.*4)

This weight is termed global because the information score is based on all the assessed seed items

A variation on this scheme allows a different set of weights to be used for each item. This is accomplished by using information scores for each item rather than the average information score:

*w_α_ (e,i) = 1_α_(statistical accuracy score)×statistical accuracy score(e) × I(f_e,i_ | g_i_)*	(*A1.*5)

For each α we define the Item weight *DM_α_* for item *i* as

*IDM_α_(i) = ∑_e=1,..E_ w_α_(e,i) f_e,i_ / ∑_e=1,..E_ w_α_(e,i)*	(*A1.*6)

The *optimized item weight DM* is *IDM_α*_* where *α** maximizes

*statistical accuracy score(IDM_a_) × information score(IDM_α_).*	(*A1.*7)

The non-optimized versions of the global and item weight DM’s are obtained simply by setting *α = 0.*

Item weights are potentially more attractive as they allow an expert to up- or down- weight him/herself for individual items according to how much (s)he feels (s)he knows about that item. "knowing less" means choosing quantiles further apart and lowering the information score for that item. Of course, good performance of item weights requires that experts can perform this up- down weighting successfully. Anecdotal evidence suggests that item weights improve over global weights as the experts receive more training in probabilistic assessment. Both item and global weights can be pithily described as optimal weights under a strictly proper scoring rule constraint. In both global and item weights statistical accuracy dominates over information, information serves to modulate between more or less equally well calibrated experts. Definitions and proofs of these scoring rule properties are found in Cooke, 1991.^10^

Since any combination of expert distributions yields assessments for the seed variables, any combination can be evaluated on the seed variables. In particular, we can compute the statistical accuracy and the information of any proposed decision maker. We should hope that the decision maker would perform better than the result of simple averaging of distributions, called the *equal weight DM*, and we should also hope that the proposed DM is not worse than the best expert in the panel. The global and item weight DM’s discussed above (optimized or not) are *Performance-based DM’s.* In general the optimized global weight DM is used, unless the optimized item weight DM is markedly superior.

The optimization in *(A1.5)* and *(A1.7)* often causes experts to be unweighted, even experts with good scores. Such experts are not “rejected;” unweighting simply means that their input is already captured by a smaller subset of experts. Their value to the whole study is brought out in studying the robustness of the optimal *DM* under loss of experts.

**Elicitation Results**

This section summarizes the structured expert judgment study of the global burden of exposure to Metals conducted by Pure Earth. The complete data on the elicitation is available on request. Table 1X gives experts’ names and affiliations. Table 2X shows the elicited metals and their abbreviations, and Table 3x their most impacted regions and most severe exposure pathways.

**Table S1: Summary of Experts and their Affiliations**

| **Expert Name** | **Expert Affiliation** |
| --- | --- |
| Dr. Roberto Bertollini | Member of the Scientific Committee on Health, Environment and Emerging Risks (SCHEER)  European Commission DG Sante |
| Dr. Patrick Breysse | Johns Hopkins’ Bloomberg School of Public Health |
| Dr. Jack Caravanos | New York University’s School of Global Public Health |
| Dr. Lillian Corra | Global Alliance on Health and Pollution |
| Dr. David Hanrahan | Pure Earth |
| Dr. Howard Hu | University of Southern California’s Keck School of Medicine |
| Dr. Amalia Laborde | Republic University of Montevideo, Uruguay |
| Dr. Ernesto Sánchez Triana | World Bank |
| Dr. Mona Wells | The Meadows Center for Water and the Environment |

**Table S2: Elicited Metals and their Abbreviations**

| **Chemical** | **Abbreviation** |
| --- | --- |
| Asbestos | ASB |
| Arsenic | AS |
| Benzene | BZ |
| Cadmium | CD |
| Chromium | CR |
| Dioxins | DF |
| Fluoride | FF |
| Highly Hazardous Pesticides | HHPs |
| Lead | PB |
| Mercury | HG |
| Polycyclic Aromatic Hydrocarbons | PAHs |
| Polychlorinated Biphenyls | PCBs |
| Polyfluorinated Substances | PFAs |
| Phthalates | FH |
| Endocrine Disrupting Chemicals | EDCs |
| Brominated Flame Retardants | BFR |

**Table S3: Summary of HICs and LMICs most impacted by chemical exposures, and their associated exposure pathways**

| **Chemical** | **High Income Countries Most Impacted** | **Low- and Middle-Income Countries Most Impacted** | **Most Severe Major Exposure Pathways** |
| --- | --- | --- | --- |
| **Asbestos** | U.S.  Canada  Australia  U.K. | Brazil  China  India  Russia | Occupational (mining)  Environmental Airborne |
| **Arsenic** | U.S.  Australia  HICs located within the Andes (e.g. Chile, Argentina) | Bangladesh  China  India  Southeast Asia (e.g. Vietnam, Cambodia) | Drinking/Groundwater  Occupational |
| **Benzene** | U.S.  OPEC Members | China  India Venezuela  OPEC Members | Occupational  Tobacco Smoke  Environmental |
| **Cadmium** | U.S.  Canada  Germany  Japan | Brazil  China Mexico | Occupational  Food  Smoking |
| **Chromium** | U.S.  Canada  Türkiye | China  India  Kazakhstan  Russia | Occupational  Drinking Water  Other: Welding, Plating, Fabrication, and Electronics Production |
| **Dioxins** | U.S.  Australia  U.K. | China  India  Southeast Asia (e.g. Vietnam, Cambodia) | Soil*  Foodstuffs |
| **Fluoride** | U.S.  Canada  Saudi Arabia  EU Countries | Bangladesh  Brazil  Mexico  Pakistan | Water  Environmental Airborne  Other: Main exposure pathways differ between HICs (water fluoridation) and LMICs (fluoride mining) |
| **HHPs** | U.S.  Canada  Eastern Mediterranean Region Countries | China  India  Indonesia  Mexico  Russia  Ukraine | Occupational (Farming)  Environmental Food |
| **Lead** | U.S.  France  Australia  U.K | Bangladesh  India Peru  Russia  Ukraine | Occupational (battery recycling)  Paint  Environmental Food/Cookpots |
| **Mercury** | U.S.  Canada  Japan  Australia | China India  South American Countries (Peru, Brazil, Chile) | Occupational (gold mining)  Environmental Food  Environmental Airborne |
| **PAHs** | U.S.  Canada  Australia  Germany  South Africa  Türkiye | China  Ghana  India  Russia | Occupational  Environmental Airborne |
| **PCBs** | U.S.  Canada  Japan  Taiwan | Brazil  China  India Central Asian Countries | Food  Occupational Airborne |
| **PFAs** | Ubiquitous** | Ubiquitous** | Water  Consumer Goods  Occupational |
| **Phthalates** | Ubiquitous** | Ubiquitous** | Water  Food |
| **EDCs** | Ubiquitous** | Ubiquitous** | Occupational  Food |
| **Brominated Flame Retardants** | U.S.  Canada  Japan  U.K. | China  India  Indonesia  Ghana | Food  Occupational |

*** - Indicates a unanimous consensus in ranking of exposure pathways**

**** - Ubiquitous exposure makes it difficult to single out countries impacted by chemicals**

A series of elicitation questions were asked of each metal,F1...F4 in addition to qualitative information, as illustrated in Table 4X for Asbestos.

**Table S4**

Example Elicitation Questions, Asbestos

| **ASB_F1** | Premature deaths*** |
| --- | --- |
| **ASB_F2** | DALYs lost*** |
| **ASB_F3** | Percentage deaths in high-income countries |
| **ASB_F4** | Percentage deaths in low- and middle- income countries |
| 1. Countries most impacted (high-income)  2. Countries most impacted (low- and middle-income)  3. Is the current level of science regarding dose-response analysis adequate?  4. Is the current level of science regarding exposure levels adequate?  5. Please rank the major exposure pathways listed here from 1 (most severe) to 5 (least severe). Please define "other" if you rank it highly | |

In addition to the variables of interest, experts also answered 17 calibration questions from their field to which true values were known, though not known by the experts at the elicitation. The calibration variables are given in Table 5X.

**Table S5**

| **Calibration Number** | **Calibration Question** |
| --- | --- |
| **CAL01** | Global emissions of Hg resulted from artisanal and small scale gold mining in 2015 |
| **CAL02** | Median estimated number of foodborne deaths due to aflatoxins |
| **CAL03** | Estimated median global foodborne DALYs due to aflatoxins |
| **CAL04** | Estimated median foodborne illness, attributed to lead, 2015, for SEAR D sub-region |
| **CAL05** | Percentage of premature deaths due to indoor pollution in India, 2019 |
| **CAL06** | Percentage of premature deaths due to indoor pollution in Bangladesh, 2019 |
| **CAL07** | Percentage of fatalities due to unintentional pesticide poisoning in America (Caribbean, Central, North and South) |
| **CAL08** | Annual estimate of underreported of suicides in India |
| **CAL09** | Average DDT in fat in adult penguins in 2008 |
| **CAL10** | Number of deaths from occupational exposure to benzene in 2019, according to IHME |
| **CAL11** | Health workers per 10,000 in France, 2019 |
| **CAL12** | Health workers per 10,000 in Sudan, 2019 |
| **CAL13** | Mean ambient particulate matter pollution in China, 2010 |
| **CAL14** | Mean ambient particulate matter pollution in India, 2010 |
| **CAL15** | Upper limit estimated number of liver cancer cases in China in people with hepatitis B virus |
| **CAL16** | Percentage change in rate premature deaths from asthma between 2010 and 2019 |
| **CAL17** | Percentage of global premature deaths from modern pollution in 2015 |

For each variable of interest and each calibration variable, experts supplied their 5%, 50% and 95% values. Two combinations of the experts’ judgments were applied, Performance Weighted (PW) and Equal Weighted (EW).

For the variables of interest, the results of these two combinations are given in Table 5X

**Table S6: Estimated Premature Deaths Lost as a Result of Chemicals, by Type**

| **Chemical** | **PW5%** | **PW50%** | **PW95%** | **EW5%** | **EW50%** | **EW95%** |
| --- | --- | --- | --- | --- | --- | --- |
| **ASBF1** | 4.82E+04 | 2.74E+05 | 3.46E+05 | 153.7 | 1.69E+05 | 6.80E+05 |
| **ASF1** | 21.71 | 1.36E+05 | 4.82E+05 | 21.26 | 3.07E+04 | 3.48E+05 |
| **BZF1** | 1.916 | 3,472 | 6,998 | 1.573 | 2,482 | 5.71E+04 |
| **CDF1** | 2.707 | 8,327 | 4.89E+04 | 2.613 | 1,878 | 5.35E+04 |
| **CRF1** | 4.449 | 5.90E+04 | 1.46E+05 | 1.195 | 1,732 | 1.07E+05 |
| **DF1** | 0.05017 | 47.89 | 325.7 | 0.05213 | 29.25 | 5.29E+05 |
| **FF1** | 0.05099 | 48.12 | 137.6 | 0.05547 | 29.34 | 7.82E+04 |
| **HHPF1** | 142.1 | 2.52E+05 | 4.84E+05 | 10.48 | 1.34E+05 | 2.38E+06 |
| **PBF1** | 944.9 | 1.66E+06 | 3.23E+06 | 690.7 | 4.85E+05 | 3.31E+06 |
| **HGF1** | 0.8269 | 7.98E+04 | 1.50E+05 | 0.2975 | 1,848 | 5.39E+05 |
| **PAHF1** | 8.828 | 8.99E+04 | 1.48E+05 | 2.8 | 5528 | 1.20E+05 |
| **PCBF1** | 0.2012 | 25.47 | 1989 | 0.2224 | 174.4 | 5.37E+05 |
| **PFAF1** | 0.211 | 9,460 | 5.00E+04 | 0.2345 | 741.7 | 4.44E+06 |
| **PHF1** | 0.05018 | 9.901 | 1.47E+04 | 0.05382 | 980.4 | 7.03E+05 |
| **EDCF1** | 0.05416 | 2.39E+04 | 4.52E+05 | 0.06004 | 5193 | 6.62E+05 |
| **BFRF1** | 0.05091 | 8,591 | 4.86E+02 | 0.03432 | 4313 | 3.61E+04 |

**Table S7: Estimated DALYs Lost as a Result of Chemicals, by Type**

| **Variable** | **PW5%** | **PW50%** | **PW95%** | **EW5%** | **EW50%** | **EW95%** |
| --- | --- | --- | --- | --- | --- | --- |
| **ASBF2** | 3.92E+5 | 4.229E+6 | 4.499E+6 | 1922 | 7.451E+5 | 7.253E+6 |
| **ASF2** | 378.9 | 9.867E+5 | 1.795E+6 | 316.8 | 2.816E+5 | 8.61E+7 |
| **BZF2** | 95.15 | 9.883E+4 | 1.498E+5 | 81.08 | 8.547E+4 | 2.338E+5 |
| **CDF2** | 98.79 | 1.941E+5 | 3.497E+5 | 74.02 | 60110 | 1.668E+6 |
| **CRF2** | 4.005E+4 | 2926000 | 3.977E+6 | 32.02 | 61050 | 1.668E+6 |
| **DF2** | 0.5971 | 900500 | 1.5E+6 | 0.5585 | 47970 | 2.05E+7 |
| **FF2** | 1431 | 4323000 | 2.458E+7 | 150.8 | 56710 | 2.156E+7 |
| **HHPF2** | 6241 | 9464000 | 1.498E+7 | 47.53 | 5795000 | 2.494E+7 |
| **PBF2** | 3.661E+4 | 40460000 | 6.963E+7 | 1.093E+4 | 20330000 | 8.468E+7 |
| **HGF2** | 1.821E+5 | 2953000 | 3.5E+6 | 742.4 | 1232000 | 2.223E+7 |
| **PAHF2** | 1.402E-09 | 6.149E-08 | 1.848E+6 | 3.483E-09 | 4634 | 1.671E+6 |
| **PCBF2** | 1.36E–09 | 2.884E+5 | 4.998E+5 | 2.021E-09 | 2452 | 1.587E+7 |
| **PFAF2** | 1.093E-09 | 1.834E+5 | 3.5E+6 | 2.419E-09 | 1959 | 3.727E+7 |
| **PHF2** | 1.371E-09 | 3988 | 5998 | 2.841E-09 | 3038 | 1.202E+5 |
| **EDCF2** | 3173 | 6.474E+5 | 4.994E+6 | 45.73 | 8839 | 2.991E+7 |
| **BFRF2** | 3032 | 1.992E+5 | 3.5E+5 | 1219 | 1.526E+5 | 6.663E+6 |

**Table S8: Estimated Percentage of Deaths in High Income Countries as a Result of Chemicals, by Type**

| **Variable** | **PW5%** | **PW50%** | **PW95%** | **EW5%** | **EW50%** | **EW95%** |
| --- | --- | --- | --- | --- | --- | --- |
| **ASBF3** | 0.2008 | 0.5672 | 0.6497 | 0.009023 | 0.2521 | 0.6213 |
| **ASF3** | 0.03018 | 0.1209 | 0.2123 | 0.02246 | 0.1052 | 0.2986 |
| **BZF3** | 0.2002 | 0.309 | 0.4781 | 0.003558 | 0.2689 | 0.5541 |
| **CDF3** | 0.003018 | 0.02046 | 0.02457 | 0.01012 | 0.2615 | 0.6967 |
| **CRF3** | 0.03009 | 0.0819 | 0.2956 | 0.01695 | 0.1505 | 0.5196 |
| **DF3** | 0.05014 | 0.1018 | 0.4487 | 0.01296 | 0.1291 | 0.6713 |
| **FF3** | 0.05019 | 0.1525 | 0.2811 | 0.002124 | 0.1183 | 0.4385 |
| **HHPF3** | 0.05021 | 0.1523 | 0.2725 | 0.01449 | 0.125 | 0.3517 |
| **PBF3** | 0.01011 | 0.05236 | 0.1392 | 0.01291 | 0.08065 | 0.346 |
| **HGF3** | 0.01248 | 0.1111 | 0.1785 | 0.01316 | 0.08972 | 0.46577 |
| **PAHF3** | 0.05022 | 0.1075 | 0.4656 | 0.0683 | 0.2407 | 0.5853 |
| **PCBF3** | 0.02202 | 0.387 | 0.7411 | 0.02398 | 0.1765 | 0.8068 |
| **PFAF3** | 1.427E-09 | 6.647E-08 | 0.7156 | 5.014E-09 | 0.3559 | 0.826 |
| **PHF3** | 0.2522 | 0.5332 | 0.8352 | 0.1204 | 0.4896 | 0.8331 |
| **EDCF3** | 0.2282 | 0.5006 | 0.7489 | 0.08604 | 0.2911 | 0.7579 |
| **BFRF3** | 0.25 | 0.5008 | 0.75 | 0.1116 | 0.5139 | 0.7726 |

**Table S9: Estimated Percentage of Deaths in Low- and Middle- Income Countries as a Result of Chemicals, by Type**

| **Variable** | **PW5%** | **PW50%** | **PW95%** | **EW5%** | **EW50%** | **EW95%** |
| --- | --- | --- | --- | --- | --- | --- |
| **ASBF4** | 0.3501 | 0.4297 | 0.7992 | 0.009133 | 0.6868 | 0.91 |
| **ASF4** | 0.7298 | 0.8785 | 0.9698 | 0.04681 | 0.8465 | 0.9717 |
| **BZF4** | 0.5077 | 0.6916 | 0.7998 | 0.003664 | 0.6632 | 0.9157 |
| **CDF4** | 0.7569 | 0.9796 | 0.997 | 0.2675 | 0.7384 | 0.9898 |
| **CRF4** | 0.6051 | 0.9173 | 0.9699 | 0.283 | 0.7977 | 0.9828 |
| **DF4** | 0.5612 | 0.8985 | 0.9499 | 0.1003 | 0.6477 | 0.9787 |
| **FF4** | 0.7074 | 0.8475 | 0.9499 | 0.03047 | 0.8375 | 0.9975 |
| **HHPF4** | 0.7104 | 0.8476 | 0.9498 | 0.08994 | 0.8209 | 0.9829 |
| **PBF4** | 0.706 | 0.8565 | 0.95 | 0.01048 | 0.8781 | 0.9857 |
| **HGF4** | 0.8058 | 0.8818 | 0.9699 | 0.3584 | 0.7589 | 0.9212 |
| **PAHF4** | 0.565 | 0.8445 | 0.9496 | 0.35844 | 0.7589 | 0.9212 |
| **PCBF4** | 0.2546 | 0.5634 | 0.9702 | 0.045 | 0.4651 | 0.9647 |
| **PFAF4** | 1.427E-09 | 6.647E-08 | 0.7156 | 5.089E-09 | 0.3649 | 0.8768 |
| **PHF4** | 0.2124 | 0.4268 | 0.745 | 0.2214 | 0.5271 | 0.8807 |
| **EDCF4** | 0.2422 | 0.4987 | 0.7578 | 0.1222 | 0.5888 | 0.8928 |
| **BFRF4** | 0.25 | 0.4992 | 0.75 | 0.2274 | 0.4861 | 0.8884 |

For the variables F1 (premature deaths), Figures 1X and 2X presents the information in Table S9 in graphic format.

**Figure S1. Premature Deaths, Equal Weighted (EW) 50% Decision Marker (DM):**

**Figure S2. Premature Deaths, Performance Weighted (PW) 50% Decision Marker (DM):**

**Expert Scoring**

The experts and decision makers are scored with respect to the 17 calibration variables. This is an unusually large number, 10 is the nominal number. As explained in Section 1 a large number of calibration variables subjects the experts to a very powerful test for statistical accuracy, often resulting in very low scores. Without removing calibration variables, we reduce the power of the statistical test to 10 effective calibration variables. This enables comparison with other studies and eliminates instability in the extreme right tail of the Chi square distribution. Scoring results with 10 effective calibration variables are shown in Tables 6-9X.

**Table S10: Scoring results with 10 effective calibration variables**

| ID | Statistical Accuracy | Mean rel info | | Un-Normalize | Relative information wrt EW | |
| --- | --- | --- | --- | --- | --- | --- |
|  |  | Total | Realizations | Weight | All var. | Calibration var. |
| exprt1 | 0.001797 | 4.05 | 2.971 | 0.00534 | 3.13 | 1.505 |
| exprt2 | 0.0355 | 1.985 | 2.447 | 0.08709 | 1.659 | 1.465 |
| exprt3 | 6.36E-08 | 2.7 | 4.789 | 0 | 2.402 | 2.621 |
| exprt4 | 3.64E-07 | 3.412 | 1.859 | 0 | 2.107 | 1.355 |
| exprt5 | 1.11E-06 | 1.919 | 3.537 | 0 | 1.808 | 2.513 |
| exprt6 | 7.16E-07 | 4.651 | 2.49 | 0 | 3.016 | 1.886 |
| exprt7 | 1.52E-06 | 3.454 | 3.344 | 5.07E-06 | 2.271 | 1.509 |
| exprt8 | 0.000299 | 4.71 | 3.014 | 0.000901 | 2.995 | 1.792 |
| exprt9 | 7.16E-07 | 4.181 | 1.679 | 0 | 2.568 | 1.246 |
| PW_10 Op | 0.6933 | 1.896 | 2.278 | 1.579 | NA | NA |
| EW_10 | 0.4866 | 0.9714 | 1.137 | 0.5535 | NA | NA |

The experts’ statistical accuracy scores range from 6.35E−8 to 3.56E−2. For experts performing this elicitation task for the first time these scores are not unusual. The two rightmost columns give the relative information of each expert with respect to the EW decision maker. They provide a yardstick for judging the level of disagreement among the experts themselves and are useful in judging robustness (see below).

**Range Graphs**

Range Graphs provide a picture of the elicitation data. Figure 3X shows the 90% confidence bands and medians (“ [----*----] ”) for all experts, for PW10 and EW for the first 13 calibration variables. The realization is given under each graph by “#”. A number of features are apparent. First the experts differ greatly in the size of their confidence bands. A band shown as “|” is too narrow to be reproduced. Without the control afforded by calibration variables, the over−confidence implied by narrow bands is not checked by reference to reality. Second, the confidence bands are often disjunct, meaning that the confidence bands do not overlap. Finally, EW is often distended relative to PW10 by a few experts with high values (e.g. variables, 3,5,6,10,12). This makes the differences in information scores intuitive.

**Figure S3: 90% Confidence Bands and Medians (“ [----*----] ”) for All Experts, for PW10 and EW for the first 13 calibration variables.**

**Robustness on items**

Robustness is gaged by eliminating a calibration variable or an expert, one at a time, recomputing the PW10 decision maker, and comparing the relative information of the “perturbed” PW10 decision maker with the original PW10 decision maker. These relative information numbers should be compared with the rightmost 2 columns of Table 6X. On this data robustness of experts was not possible as many experts gave no assessments for several variables of interest. The results for robustness on items are shown in Table 7X. The relative information scores in the rightmost two columns of Table 7X are an order of magnitude smaller than those in the rightmost two columns of Table 6X, indicating that the results are very robust against the loss of a single calibration variable.

**Table S11: Robustness on Items.**

| Bayesian Updates: no  Weights: item  DM Optimisation: yes |
| --- |
| Calibration Power: 0.6000 |

| **Number** | **Excl. Item** | **Relative Information** | | **Calibration** | **Relative information wrt original PW** | |
| --- | --- | --- | --- | --- | --- | --- |
|  |  | **Total** | **Calibr. Vbls** |  | **Total** | **Calibr. Vbls** |
| **1** | CAL01 | 1.908 | 2.391 | 0.5608 | 0.1813 | 0.2005 |
| **2** | CAL02 | 1.90 | 2.331 | 0.5608 | 0.0696 | 0.0252 |
| **3** | CAL03 | 1.573 | 2.105 | 0.6465 | 0.1407 | 0.1419 |
| **4** | CAL04 | 1.747 | 2.308 | 0.3538 | 0.175 | 0.1135 |
| **5** | CAL05 | 1.863 | 2.138 | 0.5608 | 0.2464 | 0.1578 |
| **6** | CAL06 | 1.577 | 2.157 | 0.5608 | 0.148 | 0.1811 |
| **7** | CAL07 | 1.879 | 2.147 | 0.7315 | 0.05731 | 0.08057 |
| **8** | CAL08 | 2.395 | 2.473 | 0.7315 | 0.7751 | 0.4018 |
| **9** | CAL09 | 2.254 | 2.543 | 0.8211 | 0.5743 | 0.368 |
| **10** | CAL10 | 2.272 | 2.371 | 0.621 | 0.5742 | 0.361 |
| **11** | CAL11 | 1.888 | 2.342 | 0.5608 | 0.09043 | 0.05729 |
| **12** | CAL12 | 1.92 | 2.417 | 0.8211 | 0.1215 | 0.249 |
| **13** | CAL13 | 1.916 | 2.421 | 0.7315 | 0.05549 | 0.06258 |
| **14** | CAL14 | 1.754 | 2.366 | 0.5608 | 0.1914 | 0.1311 |
| **15** | CAL15 | 2.246 | 2.343 | 0.621 | 0.601 | 0.3156 |
| **16** | CAL16 | 1.886 | 2.182 | 0.7315 | 0.05698 | 0.07891 |
| **17** | CAL17 | 1.903 | 2.264 | 0.621 | 0.1717 | 0.3003 |
| **18** | None | 1.896 | 2.278 | 0.6933 |  |  |
