## Supplementary figures and images for "Structured Expert Judgement Approach of the Health Impact of Various Chemicals and Classes of Chemicals"

### Fig S1.tiff

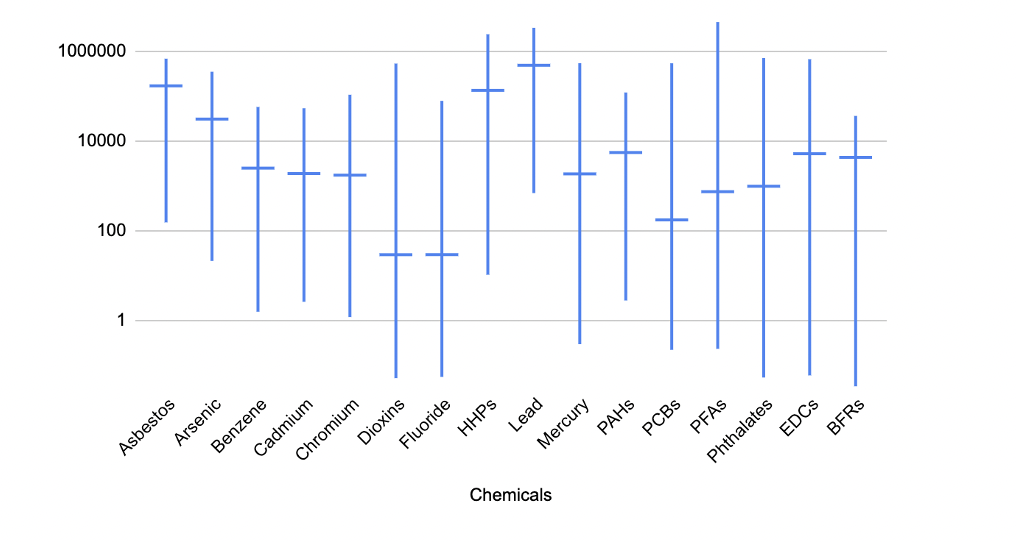

### Fig S2.tiff

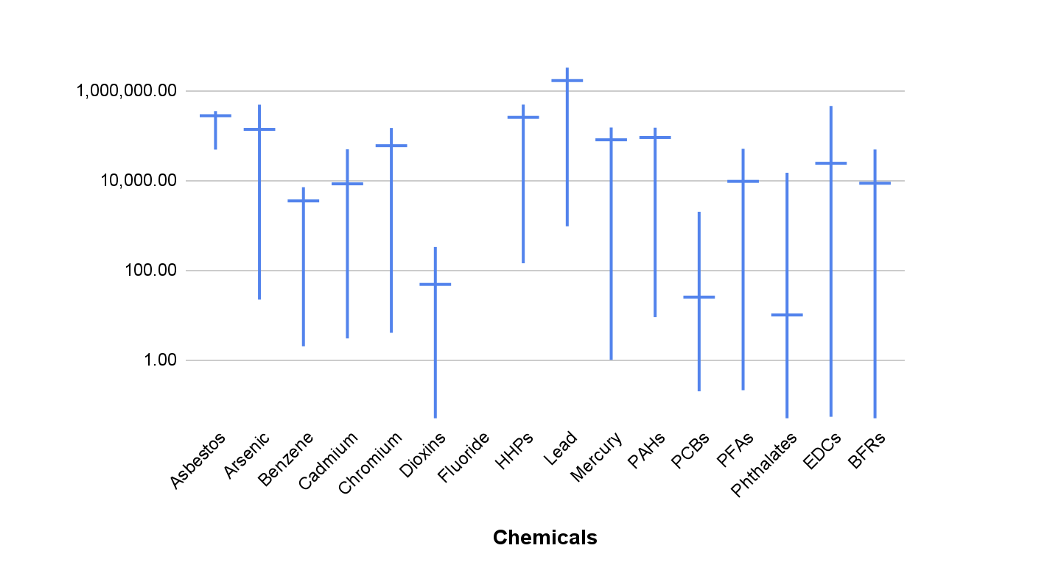

### Fig S3.tiff

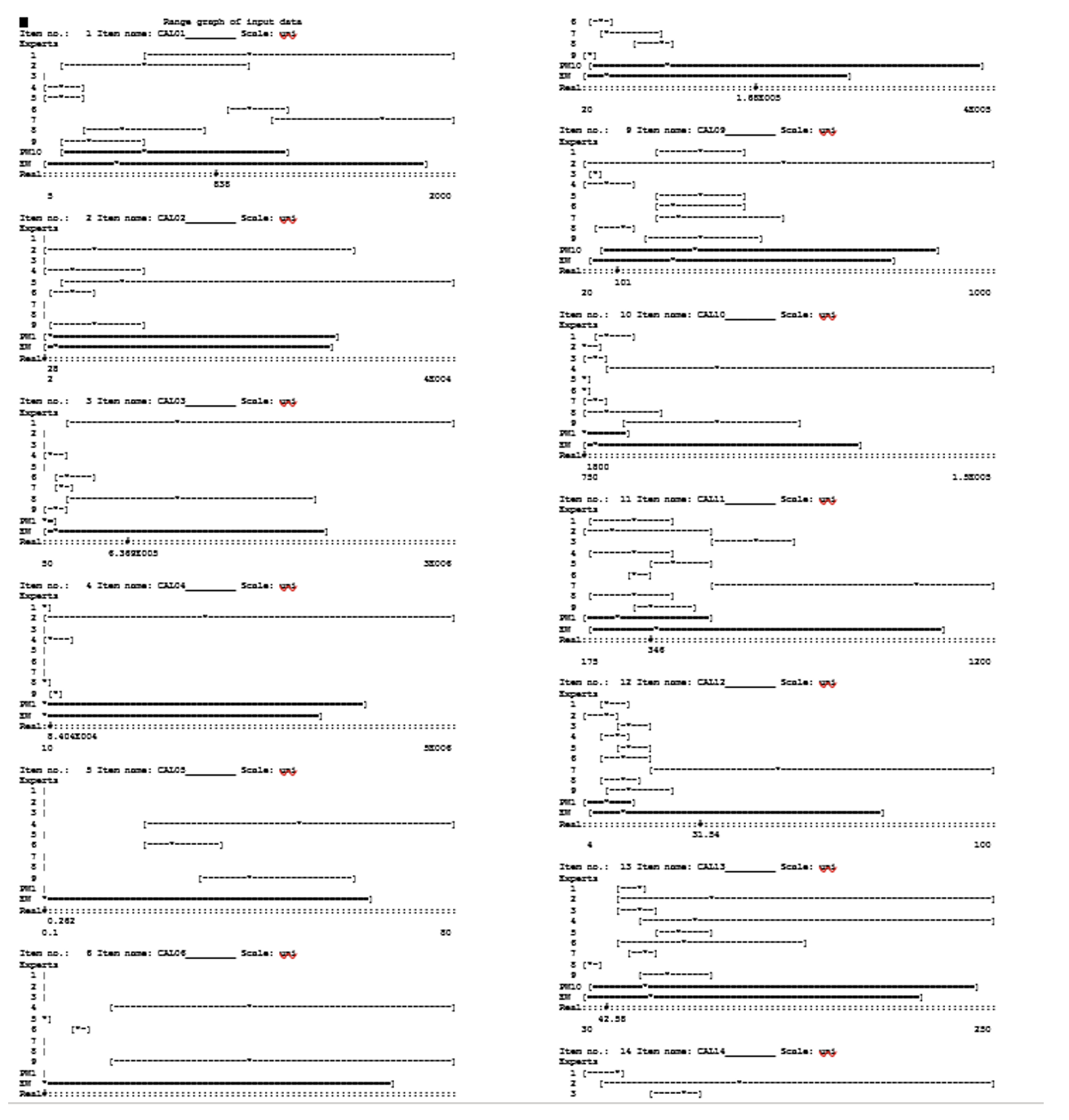
